# Through the Eyes of a Patient: Visuospatial Perspective Taking and Empathy in Medical Students

**DOI:** 10.1101/2020.04.08.20058412

**Authors:** Henryk Bukowski, Nor Faizaah Ahmad Kamal, Deirdre Bennett, Gabriella Rizzo, Colm M.P. O’Tuathaigh

**Affiliations:** Psychological Sciences Research Institute, Université Catholique de Louvain, Belgium; Medical Education Unit, School of Medicine, University College Cork, Cork, Ireland; Department of Medicine, Cork University Hospital, Cork, Ireland

**Keywords:** cognitive empathy, communication skills, perspective-taking, undergraduate medical students

## Abstract

Physicians’ cognitive empathy is associated with improved diagnosis and better patient outcomes. The relationship between self-reported and performance-based measures of cognitive empathic processes is unclear. This study examined the association between medical students’ empathy scale scores and their empathic performance in a perspective-taking task and communication skills assessment. Undergraduate medical students (N=91) completed the following: Jefferson Scale of Physician Empathy (JSPE); Empathy Quotient (EQ); a Level-1 visual perspective-taking task (VPT). Clinical communication skills were measured in a simulation-enhanced ‘breaking bad news’ assessment. Pearson’s or Spearman’s correlation coefficients and t-tests were used to determine correlation and group differences, respectively. Higher scores on the “Standing in Patients’ Shoes” sub-scale of the JSPE were associated with lower egocentric bias (*r* = -0.299, *p* < 0.05) in the VPT, which relates to capacity to adopt someone else’s perspective without being erroneously influenced by our own point of view. Additionally, a lower self-perspective advantage in the VPT, reflecting less attentional priority given to the self-perspective, was associated with higher scores on the the “Cognitive Empathy” (*r* = -0.283, *p* < 0.05) and “Emotional Empathy” (*r* = -0.342, *p* < 0.01) sub-scales of the EQ. Improved communication skills performance was associated with higher scores on the “Social Skills” EQ sub-scale (*r* = 0.298, *p* < 0.005). We show that self-assessment scores are moderately associated with performance-based measures of perspective-taking and communication skills. These results are expected to lead to improved experimental designs and a better understanding of empathy in medical education.

## INTRODUCTION

An imperative humanistic component in the development of physician-patient relationships is the physician’s ability to empathize with the patient. High levels of physician empathy are positively correlated with various indices of patient-centered care, including improved outcomes, treatment compliance, patient satisfaction, as well as a reduction in medico-legal cases (Kim et al. 2004; Hojat et al. 2011; Lelorain et al. 2018; Wang et al. 2018). Conversely, lower levels of empathy are often associated with a higher rates of physician burnout (Zenasni et al. 2012; Yuguero et al. 2017; Wilkinson et al. 2017), which may be expected to impact negatively on patient care (Gleichgerrcht & Decety, 2013).

Therefore one of the research priorities should be to design proper tools to assess physicians’ empathy. This goal is however undermined by several theoretical and methodological issues that have led researchers to observe inconsistent and inconclusive relationships between empathy scale scores and actual empathic behavior in simulated or clinical settings (Hojat et al. 2005; Chen et al. 2010; Smith et al. 2017). The first issue is that there is a lack of consensus regarding the conceptual structure of empathy and thus the way to measure empathy (Dohrenwend, 2018). In a clinical context, empathy is multidimensional, including affective, cognitive, and behavioural components (Morse et al., 1992). Affective empathy refers to the ability to experience an emotional reaction to the experience of another person, and emotional resonance is crucial in clinical settings where physicians must respond to the emotional distress of patients (Halpern 2003). The capacity to identify and understand another’s psychological perspective is referred to as cognitive empathy. The behavioural component reflects the physician’s ability to communicate and act upon that understanding of the patient’s mental condition (Morse et al. 1992; Quince et al. 2016). However, the most widely used measure of medical student empathy, the Jefferson Scale of Physician Empathy [JSPE] (Hojat et al., 2001), focuses essentially on the cognitive component of empathy, with three subscales: the “perspective-taking”, “compassionate care’ [understanding of patients’ emotions and experiences], and “standing in patients’ shoes’ [ability to see things from the patients’ perspective] (Hojat and LaNoue 2014; DiCillo et al. 2009). Another instrument frequently cited in the literature is the Interpersonal Reactivity Index (IRI; Davis 1980), which, with its “perspective taking”, “fantasy”, “empathic concern”, and “personal distress” subscales, assesses affective and cognitive components of empathy but not the behavioural component.

The second issue is that research on assessment of medical student empathy has principally relied on self-report instruments. This approach has been criticized for several reasons including poor agreement between self-report and faculty observations (O’Connor et al. 2014; Berg et al. 2015), weak correlation between self-assessed and standardized patient measures of physician empathy (Bernardo et al. 2018), as well as lack of measurement and conceptual comparability across instruments (Quince et al. 2016; Colliver et al. 2010). Hence, these issues raise justifiable concerns concerning the meaning of these scales in the context of their widespread use in medical education.

The third issue is that the instruments aimed to measure the same component of empathy actually focus on distinct aspects. For instance, cognitive empathy can be measured in terms of skills, habits, motivation, or adherence to underlying moral values. Hence, poor correlations between conceptually similar subscales across different measures e.g., “perspective-taking” across both the JSPE and IRI scales have been reported (Costa et al. 2017). Relatedly, certain authors have also noted that the distinction between the JSPE sub-scales ‘perspective-taking’ and ‘standing in the other’s shoes’ is unclear and that they may represent the same underlying factor (Stansfield et al. 2016).

The present study aimed to examine empathy among medicine students while addressing the three aforementioned issues by combining two self-report instruments and two performance-based measures, the latter including a recently developed measure of cognitive empathy which is exploited for the first time to measure medical student empathy..

Cognitive empathy is dependent upon both an unambiguous awareness of self-other distinction and the mental flexibility to adopt the subjective perspective of the other (Decety and Jackson 2004). Effective physician-patient communication is dependent upon the ability to infer the knowledge and perceptions of the patient regarding the ongoing interaction and their proximal clinical environment. It also involves the physician being able to successfully disambiguate his/her perspective on the consultation from that of the patient. These processes can be measured using visual perspective-taking tasks (VPT), where the first-person perspective is contrasted with the view of a third-person avatar, which may or may not be congruent with the research participant’s viewpoint. Perspective-taking performance in such tasks require identifying and representing another person’s visual experience, and correctly choosing the goal-relevant perspective when the self‐ and third-person perspectives are incongruent (Samson et al. 2010; Bukowski 2018). Previous studies have reported that visual perspective-taking performance is associated with increased empathy (measured using the IRI) in college students (Mattan et al. 2016; Bukowski and Samson 2017). Here, we employed an existing level-1 perspective-taking task (Samson et al. 2010; Bukowski and Samson 2017) to examine the relationship between perspective congruence and prioritization of self-vs other-person perspectives in relation with medical students’ responses in two self-report empathy measures, the JSPE (Hojat et al. 2001), and the Empathy Quotient [EQ; Lawrence et al. 2004] and observed empathy in a communication skills assessment in an objective structured clinical examination (OSCE). We hypothesized that improved VPT performance would be associated with higher empathy scores, particularly across sub-scales measuring cognitive empathic processes, in medical students.

## METHODS

### Study Design and Participants

This study was conducted in University College Cork (UCC) medical school in a cohort of Year 3 and 4 students during the first half of the academic years 2017/2018 and 2018/2019. The School of Medicine offers a systems-based integrated undergraduate curriculum with early patient contact and full time clinical placements in the latter 2.5 years of the programme. This cross-sectional study was reviewed and approved by the Clinical Research Ethics Committee of the Cork Teaching Hospitals in University College Cork.

### Instruments

#### Jefferson Scale of Physician Empathy (JSPE)

The student version of the Jefferson Scale of Physician Empathy (JSPE-S) was used to measure the empathy in medical students. It is a self-administered tool that contains 20 items across three sub-scale (“Perspective-taking”, “Compassionate Care”, and “Standing in Patients’ Shoes”) answered on a seven-point Likert-type scale (Hojat et al. 2001). The JSPE total score ranges from 20 to 140, with higher values indicating a higher degree of empathy (Kataoka et al. 2009; Magalhaes et al. 2011; Song et al. 2012; Dehning et al. 2012; Quince et al. 2016; Smith et al. 2017).

#### Empathy Quotient (EQ)

Initially designed to measure empathy in individuals with individuals who exhibit autistic traits, and validated in a clinical sample (Lawrence et al. 2004), the EQ scale is a 60-item questionnaire, where each item is a first-person statement which the study participant must rate as either “strongly agree”, “slightly agree”, “slightly disagree”, or “strongly disagree”. Previous factor-analytic studies distinguished three EQ subscales labelled “Cognitive Empathy”, “Emotional Empathy”, and “Social Skills” (Lawrence et al. 2004).

#### Level-1 Visual Perspective-taking Task

The level-1 visual perspective-taking task used in this study has been described previously (Samson et al. 2010) and correlates well with self-reported everyday life perspective-taking tendencies (Bukowski and Samson 2017). In this computer-based task, participants view pictures of a human avatar positioned in the center of a room with zero to three red discs displayed on one or two of the side walls. The avatar is viewed sideways facing either the left or the right wall. The task involves deciding whether a prompted number (ranging from 0 to 3) matches or mismatches the number of discs visible from a prompted target perspective, which could be either the participant’s perspective (self-perspective condition) or the avatar’s perspective (other-perspective condition; i.e., what number of discs are visible from the avatar’s viewpoint). The number of discs visible could be the same for both perspectives (congruent perspectives condition) or different (incongruent perspectives condition). Reaction time (RT) and error rates (ER) were collected.

The following four indices (1-4) of VPT performance were computed based on previous works (Samson et al. 2010; Bukowski 2014, 2018; Mattan et al. 2016; Bukowski and Samson 2016, 2017; Deliens et al. 2018; Bukowski et al., 2020) and two new indices (5-6) were computed. For all these indices, a higher score equates to lower VPT performance as it indicates either more biased cognition (1 to 3) or more self-centered cognition (2 and 4 to 6).

(1) The *congruency effect* reflects the extent of bias (or interference) caused by the irrelevant conflicting perspective, and captures the difficulty to handle conflicting perspectives and self-other distinction.
(2 and 3) The *egocentric* and *altercentric biases* are the extent of biases separately for self-perspective and avatar’s perspective trials, and capture, respectively, the ability to adopt someone else’s perspective without being erroneously influenced by our own point of view and the ability to evaluate one’s own perspective without being erroneously influenced by someone else’s point of view.
(4) The *self-perspective advantage* is the extent of performance advantage at judging from the self-perspective over the avatar’s perspective, and captures the attentional priority given to the self-perspective, also referred to as self-centeredness or egocentricity.
(5 and 6) The self-perspective advantage can also be computed separately for *congruent perspective trials* and for *incongruent perspective trials*. The rationale for decomposing the self-perspective advantage is that the self-perspective advantage can be expressed differently for congruent and incongruent perspectives, which respectively tap mostly on early bottom-up and later top-down processes (Samson et al. 2010; Ramsey et al. 2013).

A questionnaire was also administered which collected socio-demographic and educational/career details of the participants, such as sex, age, medical school admission pathway, nationality, and year of medical education.

#### Communication Skills Assessment

Toward the end of the 4^th^ year of medical school, students are required to take an OSCE where they are assessed on clinical skills, including their communication skills, by standardized patients. The communication skills assessment involves a breaking bad news (BBN) scenario, a challenging assignment for any doctor, requiring social and communicative competence. Several recommendations for the delivery of bad news has been established, the most well-characterised being the SPIKES protocol (Baile et al. 2000), a six-step protocol initially developed in the context of cancer care (Seifart et al. 2014). The acronym SPIKES refers to six steps recommended for breaking bad news: (a) setting up the interview; (b) assessing the patient’s perception; (c) obtaining the patient’s invitation; (d) giving knowledge and information to the patient; (e) addressing the patient’s emotions with empathic responses; (f) strategy and summary (Baile et al. 2000). A marking sheet was developed based on the student’s application of the SPIKES protocol to the BBN scenario outlined below. The assessment score range was 0-50.

The BBN assessment involved the student reviewing the case of a 36 -year-old lady who presented to her general practitioner two weeks previously with painful right eye and reduced vision, and where subsequent axial proton density–weighted MRI demonstrated multiple periventricular lesions and peripheral white matter lesions near the grey matter–white matter junction, suggestive of multiple sclerosis. The student was required to confirm to the patient the probable diagnosis of multiple sclerosis and to communicate that she needed to start treatment as soon as possible. Students had one minute to read instructions and nine minutes to complete the station.

### Procedure

The questionnaire and VPT task links were distributed via email to Year 4 medical students from January-May 2017 and January-May 2018. Briefly, all eligible students were invited to participate in a study which aimed to investigate the factors which impact on clinical communication and interpersonal understanding among medical students in clinical years. They were told that each of the online questionnaire and visual task elements would take approximately 10-15 minutes to complete. Participation was voluntary and not linked with course credits. The JSPE, EQ and socio-demographic questions were hosted online on the Typeform survey platform (https://www.typeform.com/). A link at the end of the questionnaire directed participants to the VPT task, hosted on the Testable (https://www.testable.org) behavioral testing platform. Communication skills assessments were completed by Year 4 students in April 2017 or 2018.

### Data Analysis

Summary statistical analysis was completed for categorical and non-categorical variables. Consistent with previous VPT studies (e.g. Samson et al. 2010; Bukowski 2014), only matching trials (i.e., “yes” response trials) were analyzed and Response Time (RT) was analyzed only on correct response trials. Medians were used instead of means to estimate VPT performance because all participants produced outlying RT (15% of trials on average), which excessively influences the mean but not the median (Bukowski et al. 2015). As the sample’s average accuracy was unusually low (rate of correct responses: *M* = 0.855, *SD* = 0.184), we filtered out 23 participants with outlying accuracy (i.e. a rate of accurate responses 2.5 standard deviations below the population’s median accuracy) in baseline trials (i.e., in congruent perspectives trials where no interference between perspectives could cause erroneous responses), which leaves 68 participants (rate of correct responses: *M* = 0.942, *SD* = 0.064). Analysis of variance (ANOVA) with ‘perspective’ (self-vs. avatar’s perspective) and ‘congruency’ (congruent vs. incongruent perspectives) as within-subject independent variables was conducted on correct RT and accuracy rates.

Correlational analysis using Pearson’s correlation co-efficient (*r*) was used to establish associations between (a) VPT measures and total and sub-scale scores for the JSPE and EQ; (b) communication skills scores and total and sub-scale scores for the JSPE and EQ, as well as all VPT measures. For all analyses, a *p* value < 0.05 was considered statistically significant. All statistical analyses were completed using SPSS V.20 (IBM, New York, New York, USA).

## RESULTS

### Study Demographics

91 students participated in this study, denoting a response rate of 21.9% (91/415). Age of participants ranged from 21 to 31 (M = 23.9, SD = 2.312), and 54.9% (N=50) of the sample was female. As outlined in Methods above, analysis of VPT performance data was restricted to 68 study participants.

### VPT Performance

The ANOVA revealed a non-significant main effect of ‘perspective’ (*F*(1,67) = 1.452, *p* = 0.232, η_p_^2^ = 0.021), indicating an absence of performance advantage at judging from one perspective over another, and a significant main effect of ‘congruency’ (*F*(1,67) = 38.860, *p* = 0.001, η_p_^2^ = 0.367), signifying slower RT for incongruent perspectives trials, and a significant interaction between ‘perspective’ and ‘congruency’ (*F*(1,67) = 6.164, *p* = 0.016, η_p_^2^ = 0.084). The interaction was further analyzed via pairwise t-test comparisons. The perspective advantage on congruent perspectives trials was significant (*t*(67) = 3.353, *p* = 0.001), but the perspective advantage was not significant for incongruent trials (*t*(67) = 0.698, *p* = 0.487). The egocentric bias (*M* = 87.029, *SD* = 77.873) is significantly higher than the altercentric bias (*M* = 43.723, *SD* = 138.698; *t*(67) = 2.483, *p* = 0.016).

The same ANOVA was then conducted on accuracy rates, which revealed a non-significant main effect of ‘perspective’ (*F*(1,67) = 1.329, *p* = 0.253, η_p_^2^ = 0.019), a significant main effect of ‘congruency’ (*F*(1,67) = 23.776, *p* = 0.001, η_p_^2^ = 0.262), indicating a lower accuracy for incongruent perspectives trials, and a non-significant interaction between ‘perspective’ and ‘congruency’ (*F*(1,67) = 0.881, *p* = 0.351, η_p_^2^ = 0.013).

These results replicate previous VPT studies [24, 38-40], providing confirmation that VPT performance was accurately assessed.

### Correlations between VPT Performance and Scores for the EQ and JSPE

Pearson’s correlations were computed between the six VPT indices and the total and sub-scales scores of the EQ and JSPE; results are displayed in Table 1.

**Table 1.** Correlations between the indices of visual perspective-taking (VPT) performance and the scores on the Empathy Quotient and the Jefferson Scale of Physician Empathy (JSPE).

| VPT indices | Empathy Quotient |  |  |  | JSPE |  |  |  |
| --- | --- | --- | --- | --- | --- | --- | --- | --- |
|  | Total | Cognitive | Emotional | Social Skills | Total | Patient Understanding | Compassionate Care | Standing in Patients' Shoes |
| Congruency effect | -0.142 | -0.097 | -0.147 | -0.021 | -0.095 | -0.068 | -0.022 | -0.166 |
| Egocentric bias | -0.123 | 0.048 | -0.112 | -0.159 | -0.07 | -0.036 | 0.069 | -0.299* |
| Altercentric bias | -0.138 | -0.146 | -0.171 | 0.087 | -0.107 | -0.121 | -0.074 | 0.013 |
| Self-perspective adv. | -0.314** | -0.260* | -0.321** | -0.006 | -0.054 | -0.113 | 0.062 | -0.017 |
| Self-perspective adv. for Congruent | -0.312** | -0.283* | -.0342** | 0.102 | -0.087 | -0.162 | -0.006 | 0.09 |
| Self-perspective adv. for Incongruent | -0.125 | 0.01 | -0.102 | -0.13 | 0.019 | 0.008 | 0.123 | -0.143 |

Notes: \* $p < 0.05$ ; \*\* $p < 0.01$ . Study participants were 68 medicine students.

Results indicate that less egocentric bias is associated with higher scores on the “Standing in Patients’ Shoes” sub-scale of the JSPE, and that a lower self-perspective advantage is associated with higher scores on the EQ total, as well as its “Cognitive Empathy” and, in particular, the “Emotional Empathy” sub-scale scores. Inspecting the latter VPT index separately for congruent and incongruent perspective trials reveals that the association is significant only for congruent trials.

### Correlations between communications skills assessment scores and EQ, JSPE, and VPT performance indices

Spearman’s correlations were computed between the BBN scores and the total and sub-scale scores of the EQ and JSPE; data are presented in Table 2. The results demonstrate a moderate positive correlation between communication skills performance and scores on the “Social Skills” sub-scale of the EQ. No significant correlation was observed between communications skills score and any of the six VPT task indices (all *r* ≤ 0.2, all *p* > 0.05).

**Table 2.** Correlations between communication skills performance (BBN score) and the scores on the Empathy Quotient and the Jefferson Scale of Physician Empathy (JSPE).

|  | Empathy Quotient |  |  |  | JSPE |  |  |  |
| --- | --- | --- | --- | --- | --- | --- | --- | --- |
|  | Total | Cognitive | Emotional | Social skills | Total | Patient Understanding | Compassionate Care | Standing in Patients' Shoes |
| BBN Score | 0.092 | 0.064 | 0.029 | 0.298*** | 0.119 | 0.043 | 0.009 | 0.202 |

Notes: \* $p < 0.05$ ; \*\* $p < 0.01$ ; \*\*\* $p < 0.005$ . Study participants were 91 medicine students.

### Influence of demographics

Increasing age was associated with reduced egocentric bias (*r*(68) = -0.214, p = 0.079) and self-perspective advantage for Incongruent perspectives trials (*r*(68) = -0.247, *p* = 0.042). Although VPT performance was numerically better among women across all six measures, the difference reached significance only for the global self-perspective advantage (*t*(66) = 2.185, *p* = 0.032), and for congruent perspective trials (*t*(66) = 2.780, *p* = 0.007). Females also showed higher scores for both EQ total (*t*(66) = 2.176, *p* = 0.033), and the “Emotional Empathy” EQ sub-scale (*t*(66) = 3.261, *p* = 0.002).

## DISCUSSION

This study aimed to measure for the first time in medicine students’ actual cognitive empathy performance *via* a visuospatial perspective-taking paradigm, alongside widely used self-report scales. Additionally, self-reported empathy was compared with students’ communication skills performance in an OSCE situation. Results revealed a series of meaningful associations and differences expressed on specific sub-scales and sub-components.

The first finding relates to the egocentric bias, capturing the capacity of the physician to adequately adopt the other person’s point of view when her/his own point of view may differ from those of the other person. Egocentric bias was significantly negatively correlated with only one of the self-report measures, the “Standing in Patients’ Shoes” sub-scale of the JSPE. Inspection of the two items used to score this sub-scale (“It is difficult for a doctor to view things from patients’ perspectives” and “Because people are different, it is difficult to see things from patients’ perspectives”) reveals an obvious similarity between the constructs of the egocentric bias and this JSPE sub-scale. This finding feeds into the debate whether the JSPE subscale “Perspective-taking” is adequate to capture empathic *skills* (Costa et al. 2017). Indeed, comparisons of both sub-scales reveal differences in that “Standing in Patients’ Shoes” directly asks about the physicians’ difficulties, whereas the “Perspective-taking” sub-scale asks about adherence to a set of principles and values (e.g. “A physician who is able to view things from another person’s perspective can render better care” and “Empathy is an important therapeutic factor in medical treatment.”). Hence, this finding suggests that some self-report questionnaires can (partially) capture actual performance but only items directly asking about performance might adequately do so.

The second set of findings relates to egocentricity, here measured in terms of attentional priority between the self and other perspectives while controlling (i.e., excluding) for the capacity to handle conflicting perspectives (Bukowski and Samson 2016, 2017; Deliens et al. 2018). The self-perspective advantage was significantly negatively correlated with the EQ total score, meaning that a higher EQ score predicted more other person-centered performance. Interestingly, unlike the IRI (Davis 1980) and the JSPE, the EQ contains a large portion of items directly asking about performance and only this scale showed moderate to strong correlation coefficients with our VPT measure. More importantly, the strongest association was shown between the “Emotional Empathy” subscale of the EQ and the self-perspective advantage for congruent perspective trials. The “Emotional Empathy” sub-scale differs from the “Cognitive Empathy” sub-scale in two important aspects. Firstly, the items asking about empathic performance are mainly framed in terms of experienced difficulties in Emotional empathy (e.g. “It is hard for me to see why some things upset people so much.”) and in terms of ease or competency in cognitive empathy (e.g. “I am good at predicting how someone will feel.”). Secondly, half of the items in the Emotional empathy ask about sensitivity or caring about the other person’s affective state. The sensitivity/caring dimension is conceptually close to the attentional priority given to the other person, which could explain the significant negative association with the self-perspective advantage in the VPT task. In short, the more caring/sensitivity to the other is related to a reduced self-perspective advantage, which means a higher advantage for the other person’s perspective. Congruently, a similar negative correlation between the self-perspective advantage and the ‘Perspective taking’ subscale of the IRI was recently reported (Bukowski and Samson 2017). Interestingly the items of this sub-scale ask about the extent to which one is *motivated* (and not *able*) to adopt the other person’s perspective, which also relates to caring/sensitivity for the other person.

In the current study, only the “Social Skills” sub-scale of the EQ was significantly and positively associated with communication skills performance in the BBN assessment. As described by Lawrence et al. (2004), the “Social Skills” sub-scale assesses the presence (or absence) of intuitive social skills and spontaneous and context-independent use of social skills. It has been noted that such skills are dependent upon a certain amount of cognitive empathy (Lawrence et al. 2004). Consistent with this observation, “Social skills” sub-scale scores in the current study was significantly correlated with the JSPE “Standing in the Patients’ Shoes” sub-scale (*r*_*s*_(90) = 0.397, *p* < 0.001) and the EQ “Cognitive Empathy” sub-scale (*r*_*s*_(90) = 0.439, *p <* 0.001).

Previous studies which have used self-report empathy scores show an unclear association with actual empathic behaviour. Hojat et al. (2005) reported a modest positive correlation between medical students’ self-reported empathy levels at the start of third year in medical school, and program directors’ assessments of the same students’ empathy levels during the end of internship three years later. Another study reported an apparent dissociation between self-reported empathy values and observed empathic ability, as measured in a clinical skills assessment in OSCEs completed towards the end of Years 2 and 3 of medical school (Chen et al. 2010). Specifically, while self-reported empathy declined during this time interval, more experienced students demonstrated more empathic behaviours. In a study by Smith et al. (2017), employing a longitudinal design, medical students completed self-report (including the JSPE) and behavioural (including the Reading the Mind in the Eyes Test, RMET) measures twice a year during the first three years of their studies. They demonstrated that while JSPE scores declined over this time period, the opposite pattern was observed for RMET performance, again highlighting the complex relationship between alternative measures of empathy. As mentioned in the Introduction, a large part of this heterogeneity in research findings result from (1) failing to deconstruct empathy in terms of more clearly defined components of empathy, (2) over-relying on self-report measures, and (3) a lack of critical examination regarding how a specific component is measured (e.g., motivation versus skill). In this study, we introduced a finer grained measure of cognitive empathy, the VPT, that can be further deconstructed in multiple indices of performance that differently related to motivational and skill aspects of cognitive empathy. The VPT is an interesting alternative (or complementary) tool to self-report instruments as it can be completed via internet in 12 minutes. Finally, examining correlations between the VPT indices and the specific subscales of the self-report instruments allowed to feed new critical hypotheses about the reasons why some instruments or some specific subscales fail or succeed at capturing physicians’ empathy.

One may argue that the size of the correlations reported in this study between self-reported empathy and empathy task performance are insubstantial, as they explained from 6.7% to 11.7% of the variance (*r* of 0.260 to 0.342). However, a recent meta-analysis of 85 correlational studies focused on self-report and performance-based measures of empathy showed that, on average, only 1% of the variance was explained (Murphy and Lilienfeld 2019). Additionally, a recent six-experiment study involving 1,347 participants) reported a stable correlation coefficient of 0.200 (Israelashvili et al. 2019). Furthermore, inspection of five studies that examined correlations between self-reported empathy and clinical empathic performance among medicine students reported either non-significant correlations (Haight et al. 2012), a single significant correlation (Wimmers and Stuber 2010; Berg et al. 2011; Bernardo et al. 2019), with coefficients ranging from 0.19 to 0.247, or several correlations in the range similar to the present study (LaNoue and Roter 2018).

Limitations regarding our findings include questions related to the face validity of the VPT in a healthcare context. The task does not focus on doctor-patient interactions, merely the perspectives of a non-specific avatar, but it is expected that the cognitive processes under study are pertinent irrespective of the target and VPT measures do correlates with self-reported empathic tendencies (Mattan et al. 2016; Bukowski and Samson 2017).

Another limitation is that the interpretations of the associations between VPT performance measures and specific empathy self-report sub-scales are admittedly *post hoc* and speculative and thus should be considered with caution. These findings are however fruitful in devising new ways to improve the assessment of physicians’ empathic skills.

Future studies should carefully select the self-report scales so that items directly assess specific empathy components, such as in terms of motivational states (caring about others) or performance. Subsequent research might also directly measure empathic skills with performance-based measures such as the visual perspective-taking paradigm used in the present study, especially now that some are short-lasting and internet-based. Moreover, variable outcomes arising from interventions designed to cultivate empathy in healthcare providers likely reflect the use of unidimensional summary scores which merge cognitive and affective empathy into a single concept (Martingano and Martingano 2017). In the absence of a clear conceptual definition of empathy in such types of intervention-based studies, the use of a combination of self-report and behavioural measures and/or use of multiple measures, is recommended.

## Data Availability

The data that support the findings of this study are available on request from the corresponding author, CO'T.

## Acknowledgments

We thank the students of School of Medicine, University College Cork, who helped us with the data collection, and the students who took part in our study.

